# Timing and Predictors of Tuberculosis Incidence among Contacts

**DOI:** 10.1101/2024.11.02.24316631

**Authors:** Michael Asare-Baah, Michael Lauzardo, Lori Johnston, Lina Dominique, Marie Nancy Séraphin

## Abstract

Contact investigations are crucial for tuberculosis (TB) control, yet the temporal dynamics of disease progression among exposed contacts remain poorly understood. Here, we analyzed a retrospective cohort of 44,106 contacts of 6,243 TB cases identified through the Florida Department of Health surveillance system (2009-2023). During the 15-year follow-up period, 454 individuals within this cohort developed active TB disease. Using survival analysis and mixed-effect Cox proportional hazards models, we determined that the median time to TB disease onset was 11 months following the initial contact investigation. The risk of progression to TB disease varied markedly by age and immune status. Children aged 0-5 years showed nearly seven times higher risk of TB disease progression compared to adults 65 years and older (aHR = 6.66, 95% CI: 1.33-33.27). HIV-positive contacts demonstrated a five-fold increased risk (aHR = 4.75, 95% CI: 2.43-9.30) relative to HIV-negative individuals. These findings indicate that the risk of TB transmission and disease progression persists beyond initial contact investigation activities and varies significantly across demographic and clinical subgroups, suggesting the need for extended contact monitoring and age-specific preventive strategies, particularly for young children and immunocompromised individuals.

## Introduction

Tuberculosis (TB) remains a formidable global health challenge, affecting not just those initially infected but also their close contacts. These contacts are crucial in the transmission chain and are key targets for preventive measures [1]. Individuals in close contact with active pulmonary TB cases represent a high-risk group for subsequent infection and disease development [2]. This vulnerability stems from their heightened exposure to *Mycobacterium tuberculosis* (MTB), the causative agent, transmitted via airborne droplets [3].

Contact investigations remain a crucial strategy for controlling TB disease among contacts, especially in low-incidence settings [4]. These investigations allow for the identification of both active TB disease and latent TB infection (LTBI) among exposed contacts, facilitating prompt treatment and thereby interrupting further transmission [5]. Early detection and treatment of LTBI are critical for preventing progression to active disease, while rapid identification and treatment of active TB cases minimize ongoing spread [1]. This process typically involves a comprehensive approach, including clinical assessments, chest radiography, microbiological evaluation of sputum, and tests for LTBI such as tuberculin skin tests (TST) or interferon-γ release assays (IGRA) [6],[7].

Contacts of confirmed TB cases are typically defined as individuals sharing the same airspace with an infectious individual for a significant period, often quantified as more than 15 hours per week for one or more weeks or accumulating over 180 hours during the infectious period [8]. These contacts may reside in the same household, share social settings, or be linked through workplace or school environments. Factors including the infectiousness of the index case, proximity and duration of exposure, and the inherent susceptibility of the contact influence the risk of infection among contacts [9],[6],[10].

The incidence of new TB cases among contacts following exposure is not uniform over time. It is markedly elevated during the first year post-exposure, gradually declining but remaining persistently above baseline for at least five years [6],[11],[8]. Several factors have been identified as increasing the risk of TB disease progression among contacts. These include younger age (especially children under 5), older age (individuals over 65) [12], HIV infection, inadequate antiretroviral therapy in HIV-positive individuals, a history of prior TB treatment, low socioeconomic status, certain ethnicities, comorbidities such as diabetes, specific host genetic polymorphisms, and the intensity and duration of exposure to the index case [13].

Understanding the temporal dynamics and risk factors associated with TB progression among contacts is crucial for developing targeted and effective public health interventions. By identifying those at highest risk and the critical time windows for intervention, resources can be more efficiently allocated, and preventive measures can be tailored to maximize impact. This knowledge gap forms the rationale for our study, which aims to investigate the time to TB disease onset and the specific risk factors influencing disease incidence among contacts of confirmed TB cases.

## Methods

### Study design and population

We conducted a retrospective cohort study analyzing data from the Florida Department of Health (FDOH) contact investigation database. The study population comprised 44,106 individuals identified as contacts of 6,243 index TB cases and investigated by the FDOH between January 1, 2009, and December 31, 2023. Of these contacts, 454 subsequently developed TB during the study period and were included in this analysis.

### Data Sources

We utilized two primary data sources: the FDOH Contact Investigation Database and the Florida TB Registry. The Contact Investigation Database contains comprehensive information on contacts identified, screened, and diagnosed with LTBI or active TB. The Florida TB Registry captures demographic, clinical, and epidemiological data for all reported TB cases, as documented in the Report of Verified Case of Tuberculosis (RVCT) form [14].

### Measures

#### Outcome definitions

The primary outcome of interest was the development of active TB among contacts, categorized as either incident or co-prevalent cases. We defined incident cases as contacts diagnosed with TB disease more than 30 days after the initiation of the contact investigation. Co-prevalent cases were defined as contacts diagnosed with TB disease either prior to or within 30 days of initiating the contact investigation. TB disease diagnosis was confirmed based on positive culture results for MTB or, in culture-negative cases, clinical and radiographic findings consistent with active TB as determined by a board-certified pulmonologist or infectious disease specialist. The 30-day threshold was chosen based on previous epidemiological studies [8] and accounts for the typical time required for diagnostic procedures and reporting.

#### Follow-up

We calculated the time to disease onset as the months elapsed between the contact investigation start date for the index case and the date the contact was diagnosed or reported as having TB. The follow-up period for each contact began on the date of the initial contact investigation for the corresponding index case. The endpoint was defined as either the date of TB diagnosis for contacts who developed active disease or the end of the study period (December 31, 2023) for those who remained disease-free. Time to disease onset was calculated as the interval between the contact investigation start date and the TB diagnosis date, measured in months. Contacts lost to follow-up or who died from causes unrelated to TB were censored at their last known assessment date.

#### Predictor variables

We examined a comprehensive set of risk factors for TB incidence among contacts, categorized into demographic, clinical, and social variables. Demographic factors included: Age, stratified into five groups: 0-15, 16-24, 25-44, 45-64, and ≥65 years, Sex (male/female), Birth origin (U.S.-born/non-U.S.-born), Race (White, Black, Asian), and Ethnicity (Hispanic/non-Hispanic). Clinical factors comprised: HIV co-infection status (positive/negative), determined by serological testing, Presence of drug resistance (yes/no), based on phenotypic drug susceptibility testing, and MTB strain lineage, categorized as East Asian (Lineage 2), Euro-American (Lineage 4), or Indo-Oceanic (Lineage 1), defined using whole-genome sequencing. Social factors included self-reported history within the past year for Recreational drug use (yes/no), Alcohol use (yes/no), and Homelessness (yes/no). Additional variables assessed included contact relationship to the index case (immediate family, coworker, social contact, other) and MTB clustering status (yes/no), determined by spoligotyping and 24-locus MIRU-VNTR analysis.

### Statistical Analysis

We performed a comprehensive statistical analysis to examine the characteristics and risk factors associated with TB progression among contacts. Differences in sample characteristics between incident and co-prevalent cases were assessed using Pearson’s chi-square test for categorical variables and Student’s t-test or Mann-Whitney U test for continuous variables, depending on their distribution. We conducted a time-to-event analysis to evaluate the risk of progression to active TB among contacts. Kaplan-Meier survival curves were constructed to estimate the probability of TB disease-free survival over time, stratified by key risk factors. Log-rank tests were used to compare survival distributions between groups.

To assess the individual and combined effects of various risk factors on TB incidence, we fitted univariable and multivariable mixed-effect Cox proportional hazards models. These models accounted for the clustering of contacts within index cases by including a random effect for the index case. The proportional hazards assumption was tested using Schoenfeld residuals and log-log plots [15]. Hazard ratios (HRs) with 95% confidence intervals (CIs) were reported for each predictor variable. All statistical analyses were conducted using R version 4.1.0 [16], with a two-sided α of 0.05 considered statistically significant.

## Results

### Characteristics of TB Contacts Who Progressed to Active Disease (Table 1)

Of the 454 contacts who progressed to active disease, 56.6% were co-prevalent cases, while 43.4% were incident cases. Age distribution differed significantly between co-prevalent and incident cases (p = 0.005). Most contacts were aged 45-64 years with a higher proportion of incident cases (40.1%) than co-prevalent cases (31.5%). Sex distribution was relatively balanced, with males comprising 59.0% of the total sample, and no significant difference observed between the two groups (p = 0.968). Racial distribution varied significantly (p = 0.031), with White contacts overrepresented among incident cases (55.6%) than co-prevalent cases (47.7%). Conversely, Asian contacts were more common in co-prevalent cases (9.4%) compared to incident cases (3.6%). Although not statistically significant, Hispanic ethnicity was overrepresented among co-prevalent cases (30.0%) than incident cases (23.4%) (p = 0.143). Birth origin emerged as a significant factor (p = 0.001), with incident cases showing a higher proportion of US-born contacts (70.1%) compared to co-prevalent cases (54.9%). HIV status did not differ significantly between the groups, with the majority (82.4%) being HIV-negative (p = 0.475). Drug resistance, while not reaching statistical significance (p = 0.086), was more common among incident cases (8.1%) than co-prevalent cases (3.9%). MTB lineage distribution was similar between the groups (p = 0.219), with the Euro-American lineage (L4) predominating (88.5%). There were notable differences in behavioral risk factors with recreational drug use significantly higher among incident cases (23.9%) compared to co-prevalent cases (12.8%; p = 0.003). Similarly, past-year alcohol use was more prevalent in incident cases (30.5%) than in co-prevalent cases (21.8%; p = 0.047). Past-year homelessness, while not statistically significant, was more common among incident cases (13.2%) than co-prevalent cases (8.6%; p = 0.15). Case clustering was significantly more prevalent among co-prevalent cases (92.6%) compared to incident cases (86.3%; p = 0.04). The type of contact relationship with index cases showed a trend towards significance (p = 0.063), with immediate family contacts more common among co-prevalent cases (58.0%) than incident cases (46.7%). These findings highlight important distinctions between co-prevalent and incident TB cases, particularly in terms of age, race, birthplace, substance use, and clustering patterns.

**Table 1:**
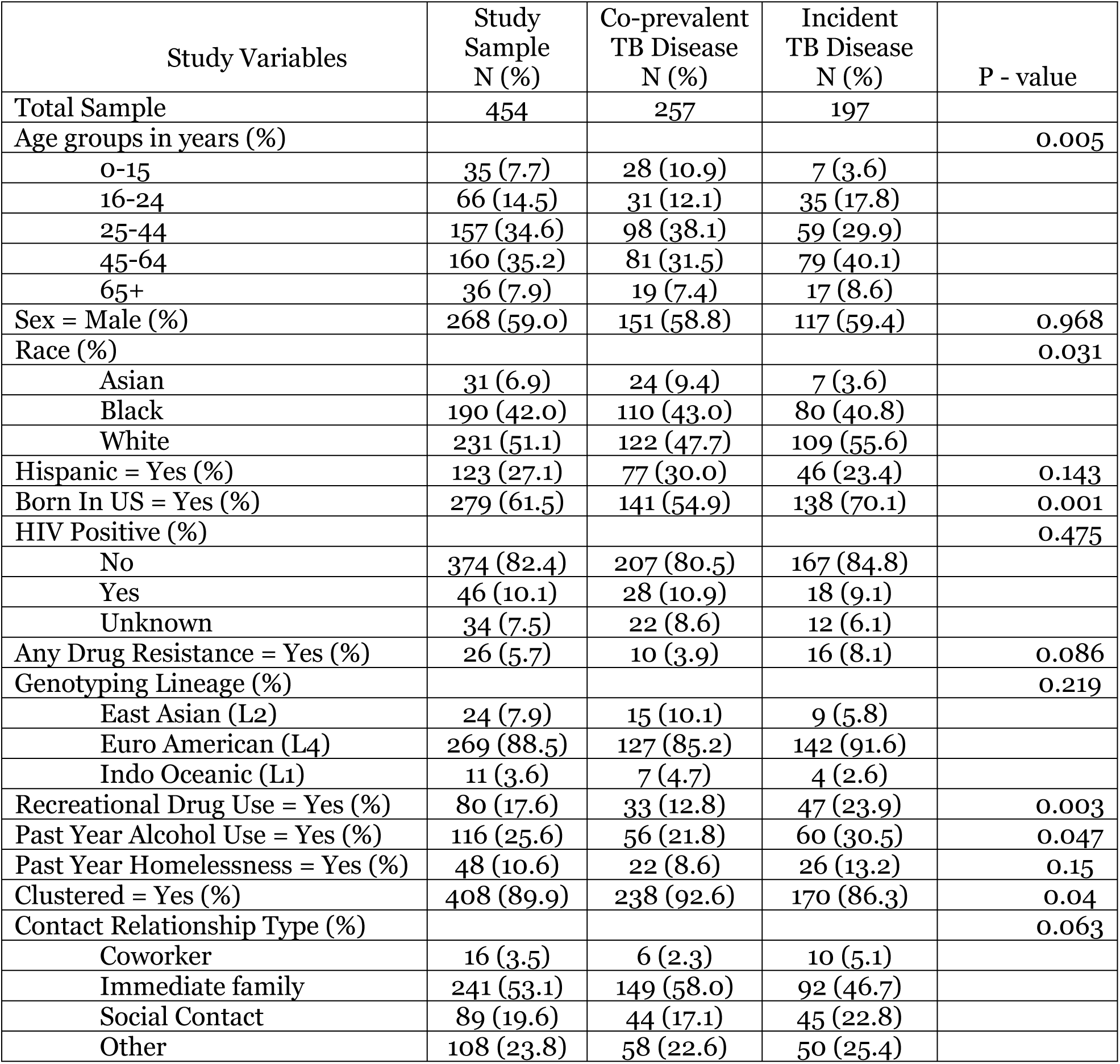
Characteristics of TB Contacts Who Progressed to Active Disease.

### Risk Factors Associated with Being an Incident TB Case Among Contacts (Table 2)

Age was significantly associated with TB disease incidence among contacts. In unadjusted analysis, children aged 0-15 years had over four times higher risk of developing TB compared to adults 65 years and older (HR = 4.21, 95% CI: 1.70-0.83, p = 0.002). This association strengthened after adjustment for confounders, with children showing nearly seven times higher risk (aHR = 6.66, 95% CI: 1.33-33.27, p = 0.021). Other age groups showed no significant difference in TB risk compared to the reference group in either unadjusted or adjusted analyses, though point estimates suggested slightly elevated risk among younger adults. HIV-positive individuals showed a substantially higher risk of developing incident TB (aHR: 4.75, 95% CI: 2.43-9.30, p = 0.0001). Interestingly, several factors often associated with TB risk in other contexts showed no significant associations in our adjusted analysis. These included sex, race, Hispanic ethnicity, US birthplace, drug resistance, recreational drug use, alcohol use, homelessness, MTB strain lineage, type of contact relationship with index cases, and contacts association with clustered cases.

**Table 2:** Risk factors associated with being an incident TB case among contacts.

| Study Variables | Unadjusted<br>HR (95% CI) | P - value | Adjusted<br>HR (95% CI) | P - value |
| --- | --- | --- | --- | --- |
| Age groups in years (%) |  |  |  |  |
| 65+ | 1.00 |  | 1.00 |  |
| 0-15 | 4.21 (1.70, 0.83) | 0.002 | 6.66 (1.33, 33.27) | 0.021 |
| 16-24 | 1.52 (1.70, 0.83) | 0.174 | 1.69 (0.81, 3.53) | 0.160 |
| 25-44 | 1.23 (1.70, 0.83) | 0.465 | 1.05 (0.54, 2.05) | 0.879 |
| 45-64 | 1.25 (1.70, 0.83) | 0.424 | 1.12 (0.57, 2.17) | 0.747 |
| Sex = Male (%) | 0.97 (0.73, 1.29) | 0.833 | 0.98 (0.66, 1.48) | 0.939 |
| Race (%) |  |  |  |  |
| Asian | 1.00 |  | 1.00 |  |
| Black | 2.29 (0.98, 0.71) | 0.056 | 3.13 (0.71, 13.90) | 0.133 |
| White | 1.65 (0.98, 0.71) | 0.242 | 2.64 (0.58, 11.95) | 0.208 |
| Hispanic = Yes (%) | 0.98 (0.70, 1.36) | 0.889 | 0.74 (0.41, 1.34) | 0.320 |
| Born In US = Yes (%) | 1.01 (0.74, 1.38) | 0.929 | 0.71 (0.44, 1.14) | 0.151 |
| HIV Positive (%) |  |  |  |  |
| No | 1.00 |  | 1.00 | 1.00 |
| Yes | 4.25 (0.76, 2.49) | 0.0001 | 4.75 (2.43, 9.30) | 0.0001 |
| Unknown | 1.37 (0.76, 2.49) | 0.296 | 1.23 (0.49, 3.09) | 0.654 |
| Any Drug Resistance = Yes (%) | 0.69 (0.40, 1.21) | 0.198 | 0.83 (0.43, 1.60) | 0.588 |
| Genotyping Lineage (%) |  |  |  |  |
| East Asian (L2) | 1.00 |  | 1.00 |  |
| Euro American (L4) | 0.95 (0.48, 0.13) | 0.875 | 0.76 (0.36, 1.59) | 0.466 |
| Indo Oceanic (L1) | 0.50 (0.48, 0.13) | 0.309 | 0.89 (0.15, 5.38) | 0.903 |
| Recreational Drug Use = Yes | 1.04 (0.75, 1.45) | 0.798 | 1.22 (0.77, 1.93) | 0.395 |
| Past Year Alcohol Use = Yes (%) | 0.85 (0.60, 1.18) | 0.326 | 0.83 (0.51, 1.35) | 0.451 |
| Past Year Homelessness = Yes | 1.38 (0.91, 2.09) | 0.135 | 1.32 (0.75, 2.32) | 0.334 |
| Clustered = Yes (%) | 1.00 (0.67, 1.51) | 0.986 | 1.22 (0.68, 2.19) | 0.511 |
| Contact Relationship Type (%) |  |  |  |  |
| Coworker | 1.00 |  | 1.00 |  |
| Immediate family | 0.84 (0.43, 0.46) | 0.598 | 0.68 (0.28, 1.64) | 0.391 |
| Social Contact | 0.80 (0.43, 0.46) | 0.523 | 0.73 (0.30, 1.78) | 0.494 |
| Other | 0.92 (0.43, 0.46) | 0.800 | 0.81 (0.34, 1.93) | 0.635 |

**Figure 1.**
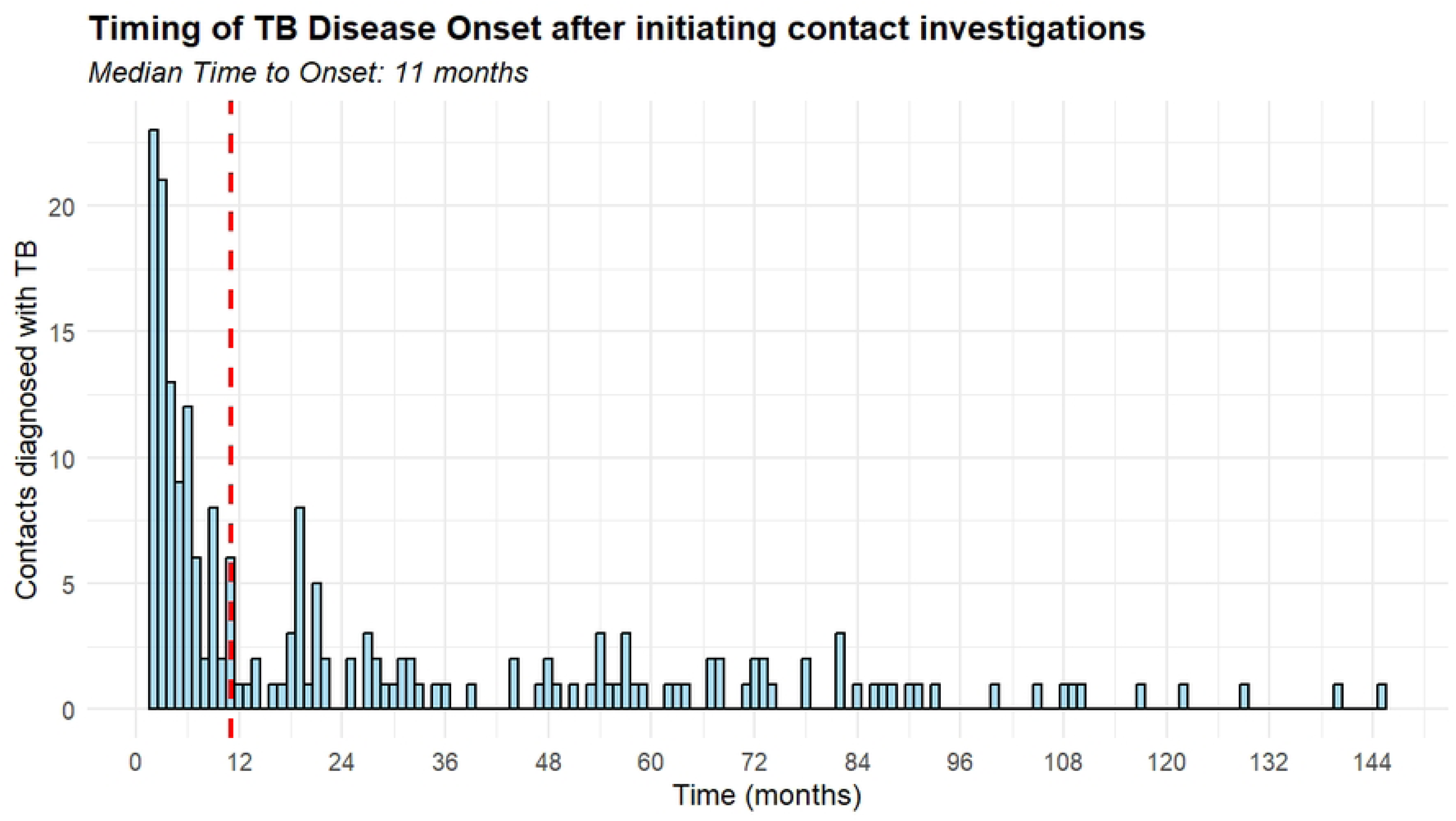
Temporal distribution of TB disease onset following contact investigation initiation. The histogram depicts the number of contacts diagnosed with TB over time (months). The Red dashed line indicates the median time to disease onset (11 months).

**Figure 2.**
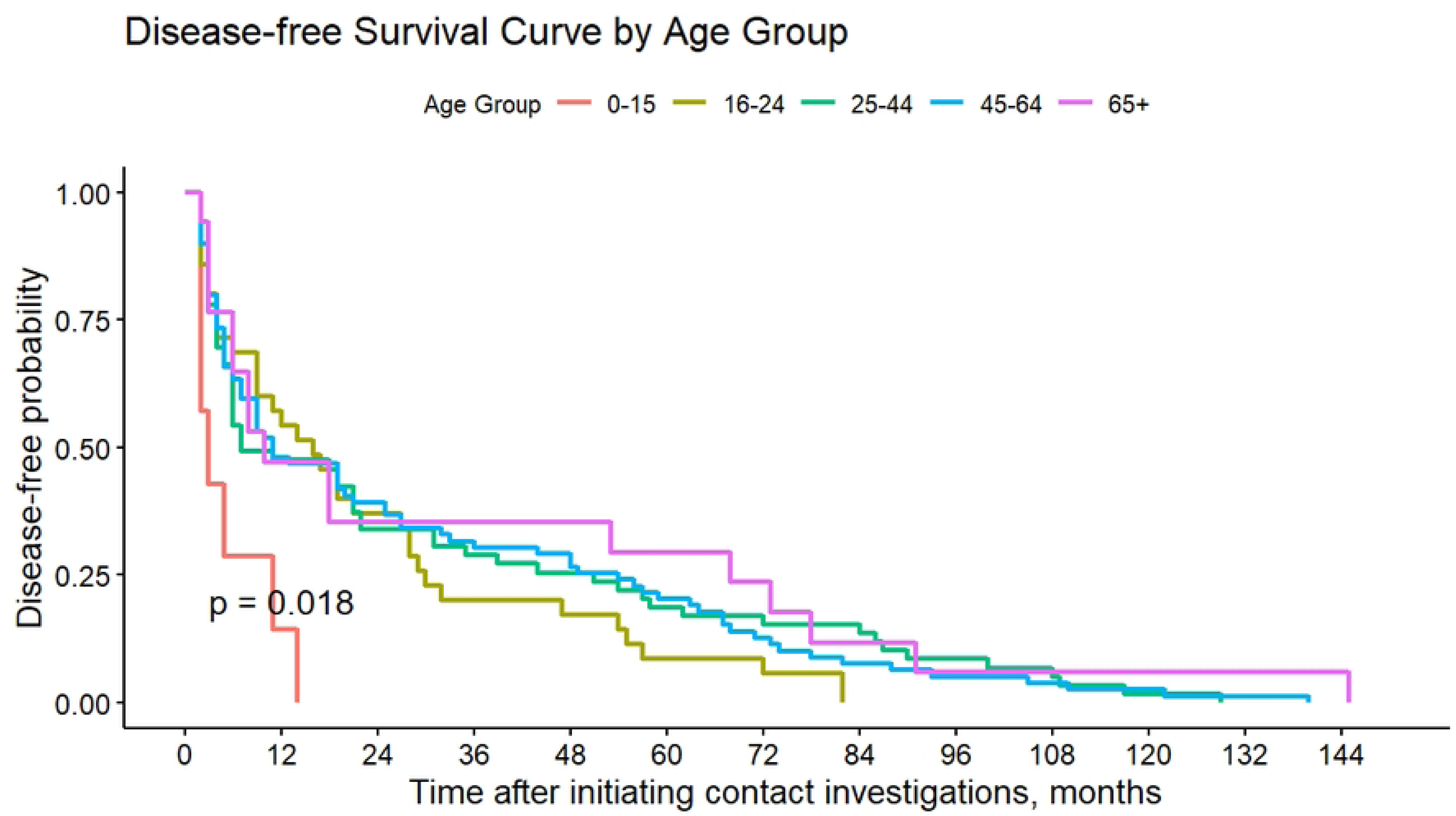
Disease-free survival curves for TB contacts stratified by age group. Log-rank test p = 0.018 indicates significant overall differences in disease-free survival across age groups with older age groups demonstrating a protective effect compared to children 0-15 years.

**Figure 3:**
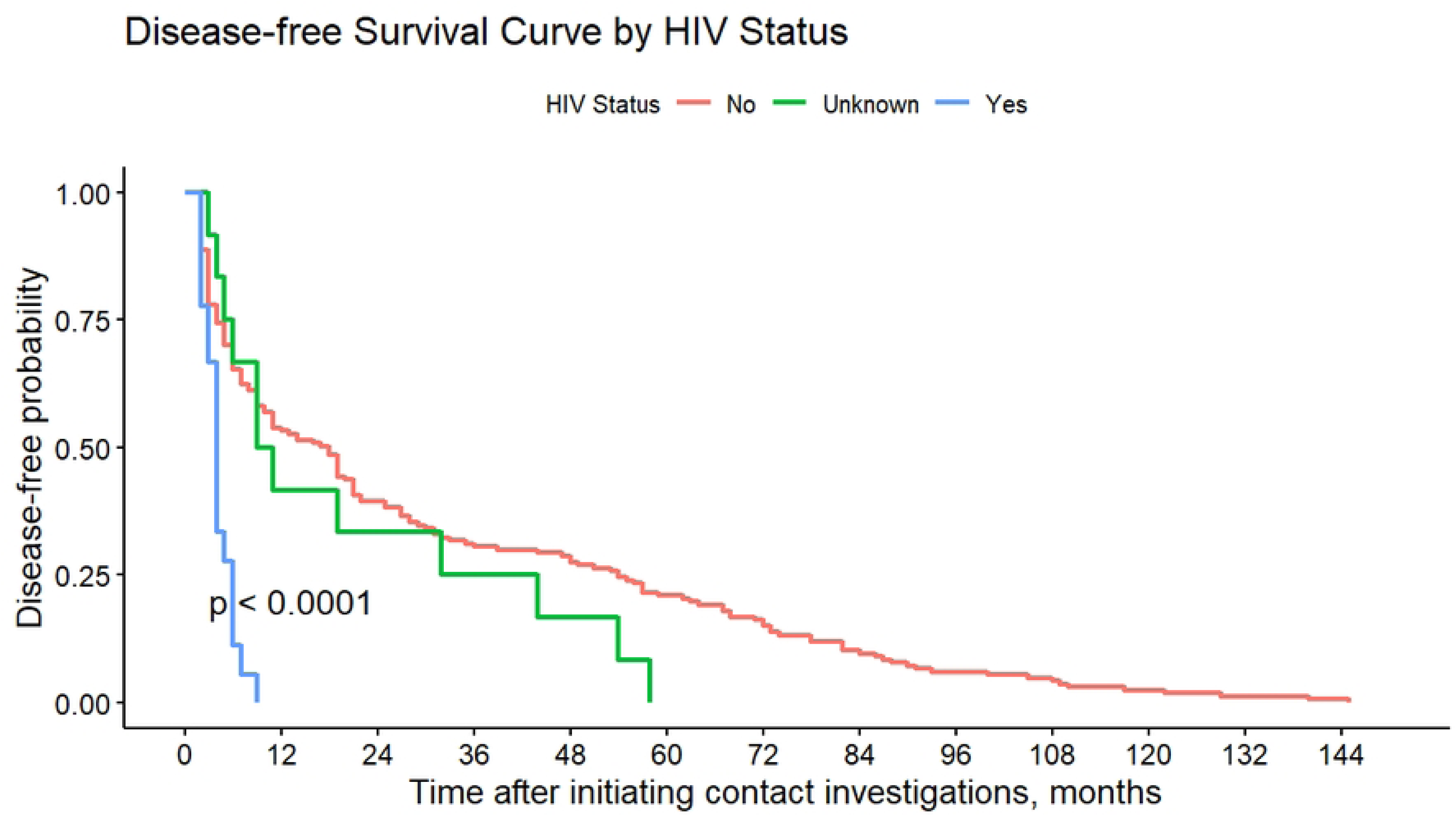
Disease-free survival curve stratified by HIV status. The graph illustrates the significant impact of HIV status on disease-free outcomes over time following the initiation of contact investigations, with HIV-positive individuals showing markedly worse disease-free probabilities, dropping rapidly within the first 12 months and approaching zero by 24 months compared to HIV-negative and unknown status groups.

## Discussion

In this statewide cohort study utilizing 15 years of data, we investigated the timing and risk factors associated with TB disease progression among contacts of confirmed TB cases in Florida. Our analysis reveals distinct patterns in disease onset and identifies key demographic and clinical factors that influence TB risk. The median time to TB disease onset of 11 months post-initiation of contact investigations observed in our study represents a critical window for surveillance and intervention. This finding aligns with previous studies that have documented similar temporal patterns in TB disease progression among contacts of infectious index cases [17],[18],[19],[5],[20]. The extended timeframe observed in the incident of TB among contacts has important implications for TB control programs. First, it reveals potential delays in diagnosis and treatment that may contribute to sustained transmission risks. Second, it demonstrates that contact susceptibility to TB infection and disease progression persists well beyond initial exposure, necessitating prolonged monitoring strategies. While the concentration of cases within the first year should guide resource allocation for TB control programs, our findings suggest that surveillance cannot be limited to the immediate post-exposure period. Extended follow-up, particularly for high-risk contacts, remains crucial for capturing late-onset cases and preventing subsequent transmission. These results emphasize the need to reevaluate current contact investigation protocols, which may need to maintain active surveillance for at least 12 months post-exposure to effectively identify and treat incident TB cases.

Our findings revealed distinct demographic and clinical features associated with TB disease risk among contacts, with age and HIV status emerging as critical determinants. Children aged 0-5 years demonstrated significantly higher susceptibility to TB compared to older age groups, consistent with previous studies [21],[22],[18]. This heightened risk likely reflects their immature immune system which is less capable of containing TB infections compared to older adults. The rapid manifestation of TB within 90 days post-exposure in children 0-5 years [23], underscores the acute vulnerability of this demographic group and the urgency for targeted preventive interventions. Interestingly, older adults (≥65 years) demonstrated lower relative risk compared to children, possibly due to broader acquired immunity and different exposure patterns. However, this finding should be interpreted cautiously, considering the limited sample size for these age groups in our study and potential confounders such as immunosenescence and comorbidities that could modify TB susceptibility. The absence of significant associations for other traditional risk factors suggests that TB progression dynamics among contacts may differ from those in the general population.

HIV co-infection emerged as another major risk factor for TB disease progression. HIV-positive contacts showed substantially increased TB risk, reflecting the impact of compromised immunity on TB susceptibility [24]. This association is particularly concerning given the elevated mortality risk observed in TB-HIV co-infected children and adolescents [25]. The challenges in accurately detecting LTBI using interferon-gamma release assays in both immunocompromised and pediatric populations further complicate early intervention efforts [26].

These results have important implications for TB control strategies. They argue for age-specific approaches, particularly intensified surveillance and prophylaxis for young children and HIV-positive contacts. Additionally, they support extending contact monitoring periods to address the prolonged window of TB disease risk, integrating active case-finding with sustained surveillance to promptly identify and treat incident cases.

## Data Availability

The dataset analyzed in this study includes sensitive patient information extracted from the Report of the Verified Case of Tuberculosis form. To protect patient privacy and maintain confidentiality, this dataset cannot be made publicly available. However, qualified researchers may access the dataset upon reasonable request, provided they adhere to appropriate data use agreements with the relevant institutional authorities that oversee the protection of this information thus the Institutional Review Board (IRB) at the Florida Department of Health (FDOH) TB Program.

## Declarations

### Human Ethics Approval and Consent to Participate Statement

This study received approval from the Institutional Review Board (IRB) at the University of Florida (approval number IRB201700445) and the Florida Department of Health (FDOH) TB Program. It was conducted as part of program evaluations and projects aimed at improving the clinical and public health services offered by state and local TB programs in Florida (SOW21-473; 6.1.11). The approval authorities waived the requirement for informed consent, as the research used existing records and did not involve direct interaction with participants.

### Consent for Publication

Not Applicable.

### Availability of Data and Materials

The dataset analyzed in this study includes sensitive patient information extracted from the Report of the Verified Case of Tuberculosis form. To protect patient privacy and maintain confidentiality, this dataset cannot be made publicly available. However, qualified researchers may access the dataset upon reasonable request, provided they adhere to appropriate data use agreements with the relevant institutional authorities that oversee the protection of this information.

### Author Contributions

MAB, MNS, and ML contributed to the study’s conceptualization and design. MAB and MNS performed statistical analyses and drafted the manuscript. LJ and LD contributed to data generation, extraction, and contextual interpretation of results. All authors critically reviewed and approved the final version of the manuscript.

## Acknowledgments

We sincerely thank the Florida Department of Health for supplying de-identified TB surveillance data used in this analysis.

## References

[1] T. D. Filardo, P.-J. Feng, R. H. Pratt, S. F. Price, and J. L. Self, “Tuberculosis — United States, 2021,” MMWR. Morb. Mortal. Wkly. Rep., vol. 71, no. 12, pp. 441–446, Mar. 2022, doi: 10.15585/mmwr.mm7112a1.

[2] C. F. Hanrahan et al., “Household-Versus Incentive-Based Contact Investigation for Tuberculosis in Rural South Africa: A Cluster-Randomized Trial,” Clin. Infect. Dis., vol. 76, no. 7, pp. 1164–1172, Apr. 2023, doi: 10.1093/CID/CIAC920.

[3] C. Greenaway, M. Palayew, and D. Menzies, “Yield of casual contact investigation by the hour,” Int. J. Tuberc. Lung Dis., vol. 7, no. 12 SUPPL. 3, 2003.

[4] Centers for Disease Control and Prevention, “Self-Study Modules 8: Contact Investigations for Tuberculosis,” 2014. Accessed: May 01, 2024. [Online]. Available: http://www.cdc.gov/tb/education/ssmodules/pdfs/module8.pdf

[5] M. R. Reichler et al., “Duration of Exposure Among Close Contacts of Patients With Infectious Tuberculosis and Risk of Latent Tuberculosis Infection,” Clin. Infect. Dis., vol. 71, no. 7, pp. 1627–1634, Oct. 2020, doi: 10.1093/CID/CIZ1044.

[6] G. J. Fox, S. E. Barry, W. J. Britton, and G. B. Marks, “Contact investigation for tuberculosis: a systematic review and meta-analysis,” Eur. Respir. J., vol. 41, no. 1, pp. 140– 156, Jan. 2013, doi: 10.1183/09031936.00070812.

[7] M. Velleca et al., “The yield of tuberculosis contact investigation in low- and middle-income settings: a systematic review and meta-analysis,” BMC Infect. Dis., vol. 21, no. 1, Dec. 2021, doi: 10.1186/S12879-021-06609-3.

[8] M. R. Reichler et al., “Risk and Timing of Tuberculosis Among Close Contacts of Persons with Infectious Tuberculosis,” J. Infect. Dis., vol. 218, no. 6, pp. 1000–1008, Aug. 2018, doi: 10.1093/INFDIS/JIY265.

[9] A. Fok, Y. Numata, M. Schulzer, and M. J. FitzGerald, “Risk factors for clustering of tuberculosis cases: A systematic review of population-based molecular epidemiology studies,” Int. J. Tuberc. Lung Dis., vol. 12, no. 5, pp. 480–492, 2008.

[10] World Health Organisation, *Global Tuberculosis Report. WHO.s.l: WHO* 2019. 2019. [Online]. Available: http://www.who.int/tb/publications/global_report/en/

[11] Y. Du et al., “Declining incidence rate of tuberculosis among close contacts in five years post-exposure: a systematic review and meta-analysis,” BMC Infect. Dis., vol. 23, no. 1, Dec. 2023, doi: 10.1186/s12879-023-08348-z.

[12] S. Godoy et al., “Risk of tuberculosis among pulmonary tuberculosis contacts: the importance of time of exposure to index cases,” Ann. Epidemiol., vol. 91, pp. 12–17, Mar. 2024, doi: 10.1016/j.annepidem.2024.01.004.

[13] P. P. Gounder et al., “Risk for Tuberculosis Disease Among Contacts with Prior Positive Tuberculin Skin Test: A retrospective Cohort Study, New York City,” J. Gen. Intern. Med., vol. 30, no. 6, pp. 742–748, Jun. 2015, doi: 10.1007/s11606-015-3180-2.

[14] CDC, “2020 Report of Verified Case of Tuberculosis (RVCT) Instruction Manual,” 2020.

[15] UCLA, “Testing the proportional hazard assumption in Cox models,” Statistical Consulting Group, 2024. https://stats.oarc.ucla.edu/other/examples/asa2/testing-the-proportional-hazard-assumption-in-cox-models/ (accessed Oct. 15, 2024).

[16] R Core Team, “R: A Language and Environment for Statistical Computing.,” R Foundation for Statistical Computing, Vienna, Austria., 2022. https://www.r-project.org/ (accessed Nov. 20, 2023).

[17] J. Singh et al., “Incidence and Prevalence of Tuberculosis among Household Contacts of Pulmonary Tuberculosis Patients in a Peri-Urban Population of South Delhi, India,” PLoS One, vol. 8, no. 7, Jul. 2013, doi: 10.1371/journal.pone.0069730.

[18] G. J. Fox, S. E. Barry, W. J. Britton, and G. B. Marks, “Contact investigation for tuberculosis: a systematic review and meta-analysis,” Eur. Respir. J., vol. 41, no. 1, pp. 140– 156, Jan. 2013, doi: 10.1183/09031936.00070812.

[19] L. Otero et al., “A prospective longitudinal study of tuberculosis among household contacts of smear-positive tuberculosis cases in Lima, Peru,” BMC Infect. Dis., vol. 16, no. 1, Jun. 2016, doi: 10.1186/s12879-016-1616-x.

[20] S. Y. Park et al., “Risk of active tuberculosis development in contacts exposed to infectious tuberculosis in congregate settings in Korea,” Sci. Rep., vol. 10, no. 1, pp. 1–10, Jan. 2020, doi: 10.1038/s41598-020-57697-1.

[21] S. Chauhan, P. Gahalaut, and A. K. Rathi, “Tuberculin skin test, chest radiography and contact screening in children ≤5 Y: Relevance in revised national tuberculosis control programme (RNTCP),” Indian J. Pediatr., vol. 80, no. 4, pp. 276–280, Apr. 2013, doi: 10.1007/s12098-012-0792-y.

[22] C. Pasqualini, L. Cohen, E. Le Roux, M. Caseris, and A. Faye, “Tuberculosis in 0–5-year-old children following TB contact investigations: a retrospective study in a low burden setting,” Front. Pediatr., vol. 11, Jul. 2023, doi: 10.3389/fped.2023.1145191.

[23] L. Martinez et al., “The risk of tuberculosis in children after close exposure: a systematic review and individual-participant meta-analysis,” Lancet, vol. 395, no. 10228, pp. 973–984, Mar. 2020, doi: 10.1016/S0140-6736(20)30166-5.

[24] N. Li et al., “Incident tuberculosis and risk factors among HIV-infected children in Tanzania,” AIDS, vol. 27, no. 8, pp. 1273–1281, May 2013, doi: 10.1097/QAD.0b013e32835ecb24.

[25] Z. Huang et al., “CRISPR detection of circulating cell-free Mycobacterium tuberculosis DNA in adults and children, including children with HIV: a molecular diagnostics study,” The Lancet Microbe, vol. 3, no. 7, pp. e482–e492, Jul. 2022, doi: 10.1016/S2666-5247(22)00087-8.

[26] A. T. Gray et al., “Treatment for radiographically active, sputum culture-negative pulmonary tuberculosis: A systematic review and meta-analysis,” PLoS One, vol. 18, no. 11 November, Nov. 2023, doi: 10.1371/journal.pone.0293535.

